# Prevalence and outcomes of recorded dementia vary by data source: a population cohort study of 133,407 older adults

**DOI:** 10.64898/2026.01.05.26343434

**Authors:** Rose S Penfold, Tim Wilkinson, Lucy E Stirland, Clare MacRae, Tom C Russ, Susan D Shenkin, Emma Vardy, Atul Anand, Bruce Guthrie, Elizabeth L Sampson, Alasdair MJ MacLullich

## Abstract

**Background:** Dementia diagnoses are captured across multiple routine data sources, but discrepancies between these may affect care and research. This study determined the prevalence and overlap of recorded dementia across primary care, hospital, and community prescribing data sources in a UK regional cohort, and examined whether outcomes differed by the setting in which dementia was first recorded.

**Methods:** Retrospective cohort study of adults ≥65 years (n=133,407) in a large Scottish health board. Dementia diagnoses recorded from 01/04/2016 to 01/04/2020 were identified across linked primary care, hospital discharge, and prescribing records. Associations between source of first recorded dementia diagnosis and subsequent mortality and emergency hospitalisation were estimated using Cox proportional hazards and Fine–Gray competing risks models.

**Results:** At baseline (01/04/2016), 7544/133407 individuals (5.7%) had recorded dementia: 95.1% in primary care, 73.3% in hospital, and 54.3% in prescribing records. Over four years, 7359 of the remaining 125,863 individuals (5.8%) had newly recorded dementia: 70.2% in primary care, 22.2% in hospital, and 7.6% in prescribing records. Only 35.9% of hospital-recorded diagnoses were coded in primary care records within a year. People first diagnosed in hospital were older, more frail, more socioeconomically deprived, and had higher mortality than those first diagnosed in primary care (<30days: adjusted Hazard Ratio (aHR) 8.96, 95%CI 6.94-13.52; >365days: aHR 1.29, 95%CI 1.19-1.41).

**Conclusions:** Dementia is variably recorded across routine datasets, and the setting in which dementia is first recorded identifies groups with markedly different prognosis. Improved data source integration and scrutiny of hospital-based diagnostic pathways are needed to ensure diagnoses are reliably transferred and people with dementia receive timely, equitable post-diagnostic care.

## Introduction

Dementia is widely recognised to be underdiagnosed and under-recorded in routine care. In the UK, around one-third of people living with dementia do not have a diagnosis in their primary care record, and studies from other high-income countries suggest up to 60% may remain undetected or undocumented in primary care.[1–7] People with dementia (diagnosed and undiagnosed) experience higher healthcare use and worse outcomes than those without.[8–12] Failure to record dementia diagnoses may deny individuals and families access to post-diagnostic support, licensed treatments and potentially new disease-modifying therapies, and advance care planning. [1, 13–16]

Electronic health records (EHR) offer new opportunities to study dementia in large populations.[17, 18] Leveraging EHR data for research can contribute to better care, for example through enhancing understanding of diagnosis rates and patterns, including how, where and when dementia is recorded, and outcomes of diagnosed dementia. However, many existing EHR-based studies have used selected cohorts and/or limited data sources, limiting generalisability.[19–23] Others have developed algorithms identifying undiagnosed dementia.[24, 25] Yet these rarely influence care, as the diagnoses are not typically accessible to clinicians across settings.

Dementia is usually diagnosed in primary care or outpatient settings in the UK and other healthcare systems.[16, 26–28] In the UK, specialist assessment is recommended for formal diagnosis, but in 2023, the average wait from referral to diagnosis was 22 weeks.[29] Meanwhile, many people with dementia are admitted to hospital, often for reasons other than dementia, at rates ranging from 0.37 to 1.26 per person-year in high-quality studies, and healthcare use often increases before a diagnosis is made.[11, 12, 30–35] Hospitalisation therefore presents a potential opportunity to recognise previously undiagnosed dementia and either make a diagnosis or initiate a post-discharge diagnostic process, complementing current outpatient-based pathways. UK guidelines support hospital-based diagnosis for appropriate cases, specifically when a diagnosis can be “made by an expert with support from multimodal assessments and collateral history, even if there is co-existing delirium”.[36–38] However, concerns persist regarding potential false positive diagnoses in hospital, particularly in the context of delirium or acute illness.[26, 39] Little is known about how many people are first diagnosed with dementia in acute hospitals and how they differ from those diagnosed in the community. The setting in which dementia is first recorded may reflect the clinical context in which dementia is recognised, and may help identify groups with different acuity, comorbidity and frailty, and with differing access to community-based memory assessment and post-diagnostic support. Understanding whether outcomes differ by diagnostic setting can help target follow-up and inform design of integrated hospital-community diagnostic and support pathways.

This study aimed to determine the prevalence and overlap of recorded dementia diagnoses across multiple linked routine data sources in a regional cohort. We further examined incident dementia over a four-year period, and its association with subsequent emergency hospitalisation and mortality, stratified by the data source of first recorded diagnosis.

## Methods

### Participants and Setting

This retrospective cohort study used routinely collected pseudonymised data accessed through the DataLoch service (dataloch.org). We included adults ≥65 years on 1 April 2016 registered with a General Practitioner (GP) in NHS Lothian (a large health board in Southeast Scotland) contributing to DataLoch. DataLoch covers ∼90% of GP practices and ∼86% of older adults in the region.[40] At least one year of GP registration was required by the index admission date.[41] The most common route to receiving a new dementia diagnosis in the community is GP referral to outpatient memory assessment services.

### Data sources

We identified dementia diagnoses using three routine data sources. General practice diagnoses were identified from Read version 2 codes. Hospital discharge data were obtained through linkage to Scottish Morbidity Records 01 (SMR01), a national administrative dataset capturing all acute inpatient care episodes, with one primary and up to five secondary diagnoses recorded using the International Classification of Diseases 10^th^ revision (ICD-10).[42] Dementia diagnoses appearing in any of the six positions were included. For both SMR01 and general practice data, dementia was identified using code lists from the Health Data Research (HDR) UK phenotype library (Appendix 1).[43] Information on prescriptions dispensed by community pharmacists was obtained through linkage to the national Prescribing Information System (PIS). Donepezil, galantamine, rivastigmine, and memantine are only recommended for NHS prescription by the Scottish Medicines Consortium for dementia treatment. Prescription of one or more of these at any time was considered specific for a dementia diagnosis.

Deaths were identified using National Records of Scotland (NRS) data, capturing all deaths in Scotland. Emergency hospitalisation data were obtained from linkage to hospital EHR (TrakCare, Intersystems), capturing all hospital admissions in NHS Lothian. Emergency admissions were identified using the admission type recorded in TrakCare.

### Other variables

The Scottish Index of Multiple Deprivation (SIMD) is an area-based measure of relative deprivation, obtained through linkage to national records.[44] The electronic frailty index (eFI) was obtained from general practice records. The eFI incorporates 36 deficits captured in routine general practice data to identify older people with frailty and has robust predictive validity for important clinical outcomes.[45] The eFI was categorised according to validated cutoffs: fit (score 0-0.12), mild (>0.12-0.24), moderate (>0.24-0.36), and severe frailty (>0.36).[45] One of the 36 deficits included in the eFI is for “memory and cognitive problems”– this includes Read version 2 codes listed in Appendix 1.

### Analyses

All analyses were performed using de-identified data within a secure data environment approved by DataLoch (DataLoch, Edinburgh, United Kingdom). Analyses and data visualisation were performed using R version 4.4.0 and packages (tidyverse, eulerr, tableone, survival, cmprsk).

All categorical variables are presented as frequencies and percentages, and continuous variables as mean (standard deviation) or median (interquartile range), as appropriate.

### Analysis: Prevalent Cohort

The prevalence of recorded dementia was determined using all three data sources on 1 April 2016. The proportion and overlap of recorded dementia diagnoses in the three data sources was graphically represented using a proportional Euler diagram showing absolute frequencies and percentages.

### Analysis: Incident Cohort

New dementia diagnoses were examined over a four-year period (1 April 2016 – 1 April 2020) by the data source in which they were first recorded. If a diagnosis appeared in multiple data sources on the same date, the first source was considered in the following order: hospital, general practice, prescribing data, with the rationale that a same-day community diagnosis likely reflects retrospective coding in the general practice records after hospital discharge.

We examined the number (proportion) of new dementia diagnoses subsequently recorded in the other two data sources at 30 days, 90 days, one year, and three years after first recorded diagnosis or at time of death, if earlier.

Cumulative incidence functions (CIF) for emergency hospitalisation and death were plotted to show risk of these events over time, accounting for competing risks. Separate curves were generated for individuals with dementia diagnoses first recorded in general practice, hospital and community prescribing data.

To examine whether the data source in which a dementia diagnosis was first recorded was associated with mortality, we used a Cox proportional hazards (PH) model, adjusted for age, sex, eFI and SIMD. A key assumption of this model is that differences between the groups remain proportional over time, and we tested this using standard tests in R and visual inspection of Schoenfeld residuals.[46] Because this assumption was not met for the first diagnosis data source variable, we used a piecewise Cox PH model, which allows the association to differ across time periods after diagnosis.[47] Based on the survival curve (Supplementary Figure 1) and clinical judgement, we used cut points at 30, 60, 180 and 365 days. Follow-up was from first recorded dementia diagnosis until death, or 23 October 2023 if still alive.

We used a Fine-Gray model to estimate the association between first diagnosis data source and risk of emergency admission, adjusted for age, sex, eFI and SIMD. This approach accounts for the fact that some people die before they can have an emergency admission. Follow-up was from first recorded diagnosis until the first emergency admission, death, or 23 October 2023 if still alive. For people who had their first dementia diagnosis recorded in hospital, time was measured from the date of discharge from that admission, and we excluded those who died during the index admission.

### Ethics

The project received approval through DataLoch, including: Caldicott Guardian approval, ACCORD sponsorship (AC23107), favourable ethics opinion under DataLoch’s delegated authority (Reference: 22/NS/0093), and a public value assessment by DataLoch’s Public Reference Group.

## Results

### Prevalent Cohort

#### Baseline Characteristics

Of 133,407 individuals ≥65 years registered with an NHS Lothian GP on 1 April 2016 (mean age 75.1 years; 55.9% female), 7544 (5.7%) had dementia recorded in one or more of the three data sources (Supplementary Table 1). Characteristics of the study population are shown in Table 1, stratified by dementia status. People with recorded dementia were on average older (mean age 83.5±7.3 years vs. 74.6±7.2, p<0.001), more likely female (65.7% vs. 55.3%, p<0.001), and had higher eFI scores (11.3% vs. 1.9% with severe frailty, p<0.001), compared to those with no recorded dementia.

**Table 1.** Baseline demographics and characteristics of 133,407 adults aged ≥65 years registered with a GP in NHS Lothian on 1 April 2016, and the subset of 7359 people with new dementia diagnoses recorded between 1 April 2016-1 April 2020, stratified by data source.

|  | Whole cohort<br>(n=133,407) |  | Incident cohort<br>(new dementia diagnoses)<br>(n=7359) |  |  |
| --- | --- | --- | --- | --- | --- |
|  | No recorded<br>dementia on<br>1 April 2016<br>(n=125,863) | Recorded<br>dementia on<br>1 April 2016<br>(n=7544) | First record in<br>general practice<br>(n=5165) | First record in<br>hospital<br>discharge data<br>(n=1634) | First record in<br>community<br>prescribing data<br>(n=560) |
| **Age in years** mean (SD) | 74.6 (7.2) | 83.5 (7.3) | 82.7 (6.8) | 85.4 (7.1) | 81.2 (6.5) |
| **Age category** years |  |  |  |  |  |
| 65-69 | 38,912 (30.9) | 302 (4.0) | 147 (2.8) | 29 (1.8) | 18 (3.2) |
| 70-74 | 32,319 (25.7) | 626 (8.3) | 531 (10.3) | 103 (6.3) | 74 (13.2) |
| 75-79 | 23,559 (18.7) | 1223 (16.2) | 971 (18.8) | 211 (12.9) | 128 (22.9) |
| 80-84 | 16,562 (13.2) | 1793 (23.8) | 1362 (26.4) | 348 (21.3) | 157 (28.0) |
| 85-89 | 9721 (7.7) | 1961 (26.0) | 1296 (25.1) | 445 (27.2) | 131 (23.4) |
| 90-94 | 3795 (3.0) | 1211 (16.1) | 682 (13.2) | 354 (21.7) | 45 (8.0) |
| 95+ | 995 (0.8) | 428 (5.7) | 176 (3.4) | 144 (8.8) | 7 (1.2) |
| **Sex: Female** N(%) | 56,273 (55.3) | 4954 (65.7) | 3201 (62.0) | 994 (60.8) | 322 (57.5) |
| **Ethnicity** |  |  |  |  |  |
| Black/Asian/Other/Not stated<sup>†</sup> | 8082 (7.1) | 283 (3.9) | 286 (5.9) | 73 (4.7) | 34 (6.5) |
| White | 105,147 (92.9) | 6961 (96.1) | 4575 (94.1) | 1484 (95.3) | 492 (93.5) |
| **SIMD Quintile\*** N(%) |  |  |  |  |  |
| 1 (most deprived) | 14,105 (11.3) | 979 (13.0) | 616 (12.0) | 236 (14.5) | 53 (9.5) |
| 2 | 25,371 (20.3) | 1567 (20.8) | 1121 (21.8) | 360 (22.1) | 102 (18.2) |
| 3 | 20,095 (16.1) | 1149 (15.3) | 700 (13.6) | 218 (13.4) | 79 (14.1) |
| 4 | 22,501 (18.0) | 1524 (20.2) | 1106 (21.5) | 367 (22.5) | 110 (19.7) |
| 5 (least deprived) | 43,073 (34.4) | 2309 (30.7) | 1601 (31.1) | 449 (27.5) | 215 (38.5) |
| *\*734 missing* |  |  |  |  |  |
| **General Practice eFI\*** N(%) |  |  |  |  |  |
| Fit (0-0.12) | 69,760 (61.5) | 1276 (16.9) | 1546 (30.3) | 275 (17.3) | 177 (33.4) |
| Mild frailty (>0.12-0.24) | 32,744 (28.9) | 3241 (43.0) | 2059 (40.5) | 684 (43.0) | 225 (42.5) |
| Moderate (>0.24-0.36) | 8761 (7.7) | 2165 (28.7) | 1080 (21.2) | 409 (25.7) | 90 (17.0) |
| Severe frailty (>0.36) | 2106 (1.9) | 853 (11.3) | 402 (7.9) | 222 (14.0) | 38 (7.2) |
| *\*12,501 missing* |  |  |  |  |  |

\*SIMD: Scottish Index of Multiple Deprivation. eFI: electronic Frailty Index.
<sup>†</sup> Categories combined into single group to avoid disclosure risks due to small numbers

#### Dementia Diagnosis by Data Source

Of the 7544 people with recorded dementia diagnoses, 7172 (95.1%) were recorded in primary care, 5526 (73.5%) in hospital and 4093 (54.3%) in community prescribing data. Overall, 1,254 (16.6%) had dementia recorded in only one source: 940 (12.5%) in primary care, 279 (3.7%) in hospital data, and a further 35 (0.5%) had dementia-specific prescribing without a coded dementia diagnosis in GP or hospital records (Figure 1).

**Figure 1.**
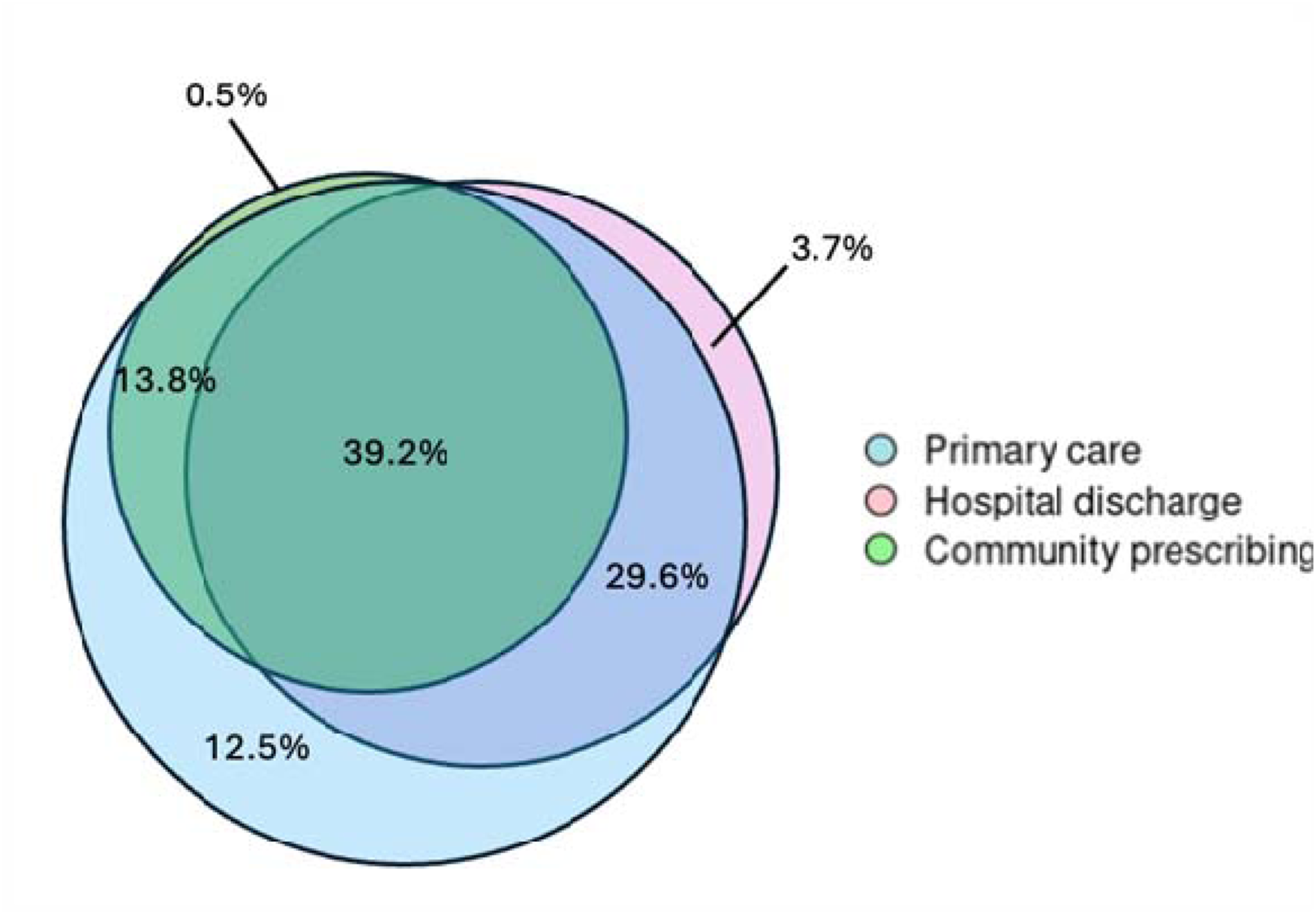
Dementia diagnoses by data source for n= 7544 adults aged ≥65 years with a recorded dementia diagnosis in one or more data sources on 1 April 2016.

#### Incident Cohort

7359 people (5.8% of the cohort with no recorded dementia at baseline) had a new recorded dementia diagnosis between 1 April 2016 to 1 April 2020. Of these, 5165 (70.2%) had their diagnosis first recorded in general practice, 1634 (22.2%) in hospital discharge, and 560 (7.6%) in community prescribing data.

Characteristics of individuals with new dementia diagnoses are shown in Table 1, stratified by data source of first recorded diagnosis. People with their first diagnosis recorded in hospital were on average older (mean age 85.4±7.1 years vs. 82.7±6.8, p<0.001), more likely living in areas of higher socioeconomic deprivation (14.5% vs. 12.0% in most deprived SIMD quintile, p<0.001), and had higher eFI scores (14.0% vs. 7.9% severely frail, p<0.001), compared to those with their first recorded dementia diagnosis in general practice.

#### Recording in Other Data Sources

The number (proportion) of new dementia diagnoses recorded in either of the two other data sources within 30 days, 90 days, one year and three years after first recorded diagnosis is shown in Table 2. Of the 1565 people discharged alive from hospital, only 562 (35.9%) had a dementia diagnosis recorded in their general practice records and 191 (12.2%) had a community prescription record within one year or at time of death, if earlier.

**Table 2.** Number (proportion) of new dementia diagnoses subsequently recorded in general practice and community prescribing data at multiple timepoints or at time of death, if earlier.

| **Data source where diagnosis first recorded** | **Time since first recorded diagnosis** | **No (%) diagnoses captured in GP data** | **No (%) diagnoses captured in community prescribing data** |
| --- | --- | --- | --- |
| GP | 30 days | - | 1158 (22.4) |
| GP | 90 days | - | 1972 (38.1) |
| GP | One year | - | 2777 (53.8) |
| GP | Three years | - | 2973 (57.6) |
| Hospital* | 30 days | 250 (16.0) | 14 (0.9) |
| Hospital | 90 days | 404 (25.8) | 75 (4.8) |
| Hospital | One year | 562 (35.9) | 191 (12.2) |
| Hospital | Three years | 625 (39.9) | 231 (14.8) |
| Community prescribing | 30 days | 73 (13.0) | - |
| Community prescribing | 90 days | 135 (24.1) | - |
| Community prescribing | One year | 203 (36.3) | - |
| Community prescribing | Three years | 240 (42.9) | - |

\*Including only those discharged alive from hospital (n=1565)

#### Outcomes

Median follow-up time following a new dementia diagnosis before death or censoring was 41.8 months (IQR 18.8-58.1). Of the 1634 people with their first dementia record in hospital discharge data, 659 (40.3%) died and 327 (20.0%) had at least one further emergency hospitalisation within a year, compared to 613 (11.9%) and 746 (14.4%) of those with their first diagnosis in general practice (Supplementary Table 2).

There were 69 people who died during the index hospital admission where dementia was diagnosed and who were excluded from competing risks analysis. Competing risks data are presented in a CIF plot (Figure 2). Mortality risk was higher at every time interval for those with their first dementia record in hospital, compared to general practice (<30days: aHR 8.96, 95%CI 6.94-13.52; >365days: aHR 1.29, 95%CI 1.19-1.41) (Table 3). The risk of emergency hospitalisation was slightly lower for those with a first diagnosis in hospital than in general practice, after adjusting for confounders and accounting for competing mortality risk (aHR 0.87, 95%CI 0.79-0.97).

**Figure 2.**
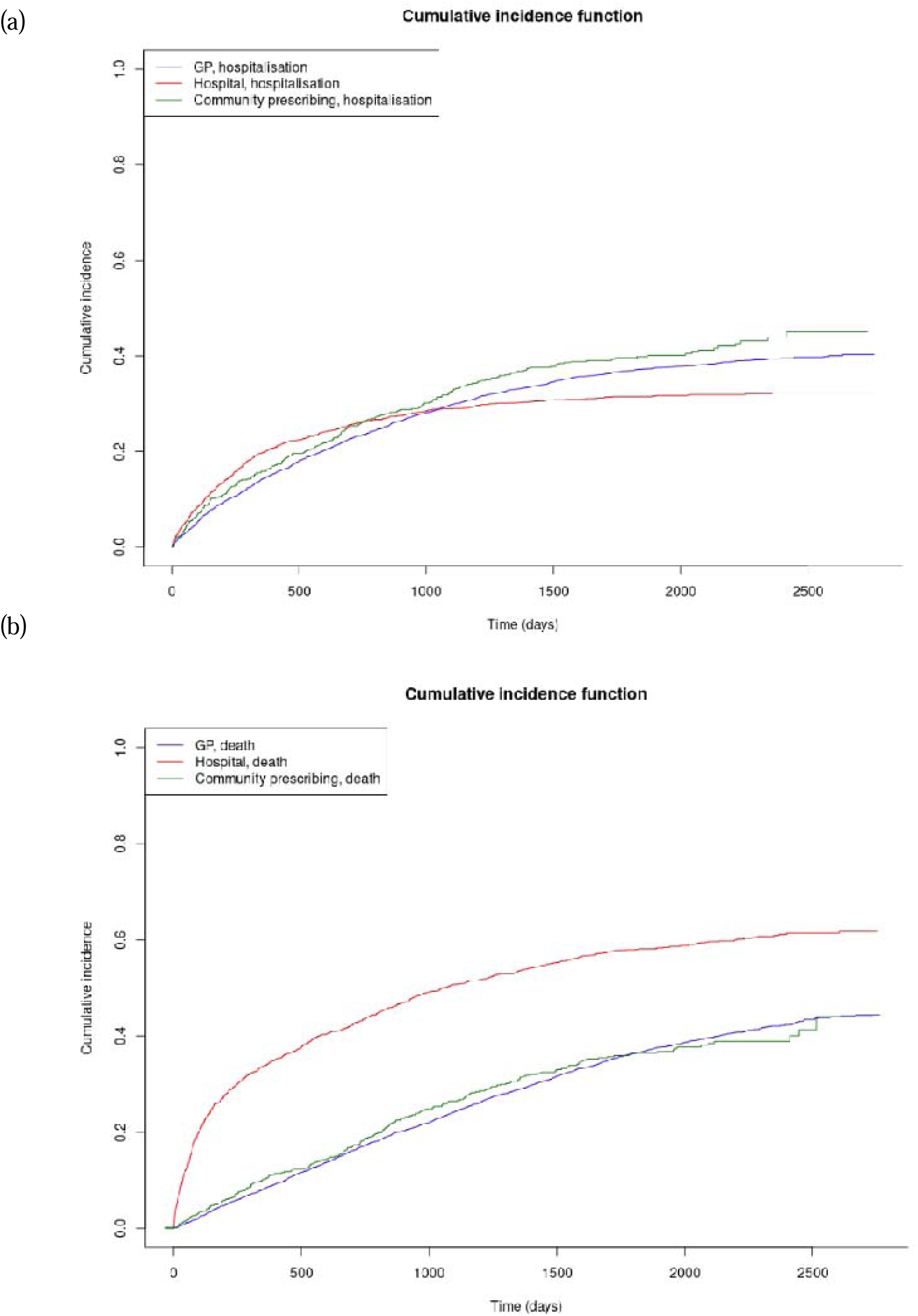
Cumulative incidence functions for competing risks of (a) hospitalisation and (b) death up to 23 October 2023 for n=7359 individuals with new dementia diagnoses between 1 April 2016 and 1 April 2020, stratified by data source where dementia first recorded.

**Table 3.** Unadjusted and adjusted hazard ratios for mortality and unplanned hospitalisation following a new dementia diagnosis between 1 April 2016 and 1 April 2020.

|  | Unadjusted HR<br>(95% CI) | Adjusted HR*<br>(95% CI) |
| --- | --- | --- |
| **MORTALITY<sup>†</sup>** |  |  |
| **Within 30 days** |  |  |
| First dementia record in primary care | Ref | Ref |
| First dementia record in hospital discharge | 11.12 (7.39-16.74) | 8.96 (6.94-13.52) |
| First dementia record in community prescribing | 1.54 (0.60-3.97) | 1.75 (0.68-4.51) |
| **Between 30-60 days** |  |  |
| First dementia record in primary care | Ref | Ref |
| First dementia record in hospital discharge | 7.73 (5.29-11.31) | 6.66 (4.53-9.78) |
| First dementia record in community prescribing | 1.43 (0.60-3.37) | 1.39 (0.55-3.53) |
| **Between 60-180days** |  |  |
| First dementia record in primary care | Ref | Ref |
| First dementia record in hospital discharge | 4.28 (3.56-5.15) | 3.56 (2.96-4.28) |
| First dementia record in community prescribing | 1.11 (0.74-1.67) | 1.27 (0.84-1.91) |
| **Between 180-365days** |  |  |
| First dementia record in primary care | Ref | Ref |
| First dementia record in hospital discharge | 2.46 (2.04-2.95) | 2.03 (1.68-2.44) |
| First dementia record in community prescribing | 1.05 (0.74-1.48) | 1.17 (0.82-1.65) |
| **After 365days** |  |  |
| First dementia record in primary care | Ref | Ref |
| First dementia record in hospital discharge | 1.54 (1.42-1.68) | 1.29 (1.19-1.41) |
| First dementia record in community prescribing | 1.00 (0.88-1.12) | 1.09 (0.96-1.23) |
| **EMERGENCY HOSPITALISATION<sup>†</sup>** |  |  |
| First dementia record in primary care | Ref | Ref |
| First dementia record in hospital discharge | 0.91 (0.82-1.00) | 0.87 (0.79-0.97) |
| First dementia record in community prescribing | 1.13 (0.98-1.29) | 1.11 (0.96-1.27) |

\* Adjusted for age, sex, SIMD, eFI
† For mortality, HR is estimated using Cox proportional hazards model; for emergency admission HR shown is the subdistribution hazard ratio from a Fine-Gray model

## Discussion

In this UK regional cohort of 133,407 people aged ≥65 years, baseline recorded dementia prevalence was 5.7%, with 16.6% of diagnoses in a single data source. Over four years, a further 7359 people (5.8% of those without dementia at baseline) had a new dementia diagnosis, with one in five (22.2%) first recorded in hospital data. Individuals with dementia first recorded in hospital were older, more frail, more socioeconomically deprived, and had higher mortality than those first diagnosed in the community.

This study is, to our knowledge, novel in combining general practice, hospital discharge, and community prescribing data longitudinally to explore routine dementia coding and outcomes in a large UK population. Previous studies have been cross-sectional, in selected cohorts, or used limited data sources.[48–51] Although it is not unexpected that previously unrecognised dementia may be identified during an acute admission, the key finding was that a substantial proportion of people had their first recorded diagnosis in hospital data, and only 36% were subsequently coded in general practice within one year despite post-diagnostic care being predominantly community-based.[26, 48, 52]. This pattern may reflect delays in receiving a definitive community diagnosis, including waiting for memory assessment services, alongside incomplete transfer of diagnostic information and variation in coding practices between settings. in the UK, liaison teams may recommend GP referral to memory services after discharge, so waiting times and imperfect communication or coding may contribute to the observed mismatch. Confirming these mechanisms would require linkage to referral/ memory service data and case notes, which were not available.

No comparable studies have examined the contribution of hospital-based diagnoses to new dementia diagnoses in a population cohort, despite UK guidelines recommending diagnosis in acute hospital settings for appropriate cases.[36, 37] A previous study in a subset of this population (UK Biobank participants living in Edinburgh) reported high positive predictive values (PPVs) for dementia recorded in both general practice (86.8%) and hospital admissions data (87.3%) compared to expert adjudication of medical records.[49] Another study of English hospital data reported a specificity of 92% compared to memory clinic diagnoses, noting this may be an underestimate, as the characteristics of patients with hospital diagnoses suggested some may be accurate diagnoses missed in memory clinic.[53] However, concerns persist regarding false positive diagnoses in hospital, particularly in the context of delirium or acute illness.[26, 39] Conversely, caution about diagnosing dementia in hospital may contribute to false negatives, if dementia is not recorded or follow-up is not initiated. Hospital-based cognitive assessments, including tools such as the 4AT, may support identification and onward referral for dementia assessment.[54] Taken together, findings highlight need for consensus on appropriate hospital-based diagnostic and follow-up pathways for people with new dementia diagnoses, including effective information transfer to general practice records.

Only 36% of hospital-based dementia diagnoses were subsequently recorded in general practice, and 12% received community prescriptions for dementia-specific medications within a year. Case notes review is needed to determine whether diagnoses were reviewed in the community, whether there was clinical disagreement, or whether this reflects gaps in coding and communication. Nevertheless, these low figures suggest potential missed opportunities for post-diagnostic support, pharmacological treatment (where appropriate), and advanced care planning, all predominantly delivered in the community.[13, 14] Lack of concordance between care settings is not unique to dementia and has been reported for other conditions diagnosed in specialist or hospital settings, including stroke and cancer.[55, 56] For dementia, the gradual onset of symptoms and diagnostic uncertainty could also contribute to delays in definitive coding in general practice and use of non-specific cognitive symptom codes. The characteristics and poorer prognosis of people diagnosed in hospital may reflect a combination of more advanced dementia and greater comorbidity and frailty at time of diagnosis, emphasising the importance of integrated, timely care across hospital and community services.

This study is an important contribution to understanding recorded dementia prevalence in a large UK region. The recorded cross-sectional prevalence in over-65s (5.7%) was only slightly lower than the 7.1% estimated in a UK-based modelling study and the 6.5% reported by Cognitive Function and Ageing Study II using 2011 data.[57, 58] In contrast, a systematic review comparing recorded dementia diagnoses in single data sources to expert-led reference standards reported widely varying sensitivities, ranging from 21%-86%, with only 3/27 studies reporting sensitivities above 60%.[19] This supports the value of multiple data sources for better ascertainment, although dementia remains underdiagnosed and/or under recorded in this population. We also show that people with dementia identified in hospital datasets are older, more frail, and more socioeconomically deprived than the wider dementia population, meaning studies relying only on hospital data risk introducing ascertainment bias.

Building on previous studies demonstrating a poor prognosis for people with dementia in hospital settings,[8, 59–62] we found that mortality is higher for those first diagnosed in hospital than for those first diagnosed in the community, even after adjustment for age, sex, frailty, and deprivation. Notably, 40% of those with hospital-based diagnoses died within one year, and the markedly elevated mortality risk in the first 30 days likely reflects the clinical context in which dementia is recognised, including acute severe illness, and high comorbidity and frailty. These associations should therefore be interpreted as prognostic differences related to the context of first diagnosis, rather than causal effects of diagnostic setting. Clinically, a first dementia diagnosis in hospital identifies a high-risk group who may benefit from proactive discharge planning, medication review, linkage to community dementia support, and advance care planning. While it is uncertain whether such interventions could improve prognosis, they may improve support and quality of life for people with dementia and their carers. The persistently increased relative mortality risk beyond one year suggests that those who survive their acute illness remain vulnerable and may benefit from ongoing post-diagnostic support and care planning. These findings also highlight that studies using a single data source may preferentially capture particular patient groups, with implications for case ascertainment and evaluation of dementia care pathways.

### Strengths and Limitations

Study strengths include the large, population-based cohort, robust individual-level linkage across general practice, hospital, and community prescribing data, longitudinal analysis to capture new dementia diagnoses and outcomes, and follow up within a national health system.

We note several limitations. We could not validate dementia cases in any data source against clinical notes or review information used by coders to generate a diagnostic code. However, a previous validation study using general practice and hospital discharge data from this region reported high PPVs of both data sources against reference standard assessment (albeit in a younger, selected UK Biobank cohort), and community prescribing data is also very likely to have high PPV.[49] Outpatient memory clinic data is not diagnostically coded in Scotland, but this information is sent directly to the GP for ongoing management including prescription of dementia-specific medications. Hence, outpatient dementia diagnoses should be captured in GP and (for appropriate cases) community prescribing data. We analysed all-cause mortality by linkage to date of death, and therefore could not determine how often dementia was recorded as a primary or contributing cause of death. Residual confounding is possible.

However, we adjusted for important covariates including a widely-validated frailty index available in routine GP data. Furthermore, the aim of the multivariable models was not to draw causal inference but to understand the prognostic effect of dementia, adjusting for key routinely available risk factors.

### Implications for Clinical Practice

This study highlights the importance of effective information transfer to ensure dementia diagnoses are available at point of care. We found that a significant proportion of people had their first dementia diagnosis recorded in hospital, yet many of these diagnoses were not subsequently coded in general practice. Consensus is needed on hospital-based pathways, specifying when a diagnosis can be made in hospital and when referral to community memory services is required for confirmation, with defined roles for GPs in coordinating follow-up and initiating post-diagnostic care. If implemented effectively, complementary hospital-based diagnosis could reduce burden on community memory assessment services, widen access and improve overall dementia diagnosis rates. There is need to understand inconsistencies between data sources, and greater standardisation of hospital discharge summaries and coding practices may further improve care continuity.

### Implications for Research

Researchers should use multiple data sources to improve sensitivity of dementia ascertainment and mitigate biases in single-source data. Accurate recording in EHRs is essential for monitoring and prevalence modelling, relevant for policy and service planning. In Scotland, it facilitates monitoring of the Scottish Government’s commitment to one year of post-diagnostic support following a new dementia diagnosis.[13] Future studies should explore hospital-based dementia diagnosis pathways by reviewing full-text clinical notes linked to coded discharge data. There is need to better characterise people diagnosed with dementia in hospital and understand whether diagnostic opportunities are being missed in the community.

## Conclusion

One in five people had their first dementia diagnosis recorded in hospital, and many of these diagnoses were not transferred to general practice records. People diagnosed in hospital were older, more frail, more socioeconomically deprived, and had higher mortality than those diagnosed in general practice. Improved data source integration and scrutiny of hospital-based diagnostic pathways are needed to ensure diagnoses are reliably transferred and people with dementia receive timely, equitable post-diagnostic care.

## Data Availability

Data may be accessed through DataLoch (dataloch.org) following successful application and approvals. Analysis scripts for this study are available upon reasonable request to the corresponding author, and following appropriate approvals from DataLoch.

## Acknowledgements

This work uses data provided by patients and collected by the NHS as part of their care and support. This project has been facilitated by the DataLoch service (reference: DL_2023_012). DataLoch enables access to de-identified extracts of health care data from the South-East Scotland region to approved applicants: dataloch.org.

For the purpose of open access, the author has applied a CC-BY public copyright licence to any Author Accepted Manuscript version arising from this submission.

