## Supplementary Materials for "Prevalence and outcomes of recorded dementia vary by data source: a population cohort study of 133,407 older adults"

**Supplementary Tables and Figures**

**Supplementary Table 1.** Recorded dementia diagnoses by data source for n=7544 adults aged ≥65 years with a recorded dementia diagnosis registered with a DataLoch GP on 1 April 2016.

| **Data Source** | **Total no. recorded in each data source** | **No. recorded only in this data source** | **Data source plus general practice** | **Data source plus hospital discharge** | **Data source plus community prescribing** | **All three data sources** |
| --- | --- | --- | --- | --- | --- | --- |
| General practice | 7172 (95.1%) | 940 (12.5%) | - | 2232 (29.6%) | 1043 (13.8%) | 2957 (39.2%) |
| Hospital discharge data | 5526 (73.3%) | 279 (3.7%) | 2232 (29.6%) | - | 58 (0.8%) |  |
| Community prescribing data | 4093 (54.3%) | 35 (0.5%) | 1043 (13.8%) | 58 (0.8%) | - |  |

**Supplementary Table 2.** Outcomes of adults aged ≥65 years with new recorded dementia diagnoses between 1 April 2016- 1 April 2020, stratified by data source where dementia diagnosis first recorded.

|  | **All new dementia diagnoses**  **(n=7359)** | **First record in general practice (n=5165)** | **First record in hospital discharge (n=1634)*** | **First record in community prescribing (n=560)** |
| --- | --- | --- | --- | --- |
| **MORTALITY** N(%) |  |  |  |  |
| Within 30 days | 190 (2.6) | 34 (0.7) | 150 (9.2) | 6 (1.1) |
| Within one year | 1346 (18.3) | 613 (11.9) | 659 (40.3) | 74 (13.2) |
| Within three years | 3325 (45.2) | 1981 (38.3) | 1117 (68.4) | 227 (40.5) |
| Before end of study period (23^rd^ October 2023) | 5065 (68.8) | 3332 (64.5) | 1374 (84.1) | 359 (64.1) |
| **EMERGENCY HOSPITALISATION** N(%) |  |  |  |  |
| Within 30 days | 178 (2.4) | 106 (2.1) | 59 (3.6) | 13 (2.3) |
| Within one year | 1162 (15.8) | 746 (14.4) | 327 (20.0) | 89 (15.9) |
| Within three years | 2189 (29.7) | 1535 (29.7) | 472 (28.9) | 182 (32.5) |
| Before end of study period (23^rd^ October 2023) | 2688 (36.5) | 1941 (37.6) | 517 (31.6) | 230 (41.1) |
| **NUMBER OF EMERGENCY HOSPITALISATIONS WITHIN ONE YEAR OF DIAGNOSIS** |  |  |  |  |
| 0 | 6195 (84.2) | 4418 (85.5) | 1307 (80.0) | 470 (83.9) |
| 1 | 953 (13.0) | 619 (12.0) | 262 (16.0) | 72 (12.9) |
| 2+ | 209 (2.8) | 127 (2.5) | 65 (4.0) | 17 (3.0) |
| **NUMBER OF EMERGENCY**  **HOSPITALISATIONS WITHIN THREE YEARS OF DIAGNOSIS** |  |  |  |  |
| 0 | 5168 (70.2) | 3629 (70.3) | 1162 (71.1) | 377 (67.3) |
| 1 | 1599 (21.7) | 1133 (21.9) | 327 (20.0) | 139 (24.8) |
| 2+ | 590 (8.0) | 402 (7.8) | 145 (8.9) | 43 (7.7) |

*includes 69 people who died during index hospital admission where dementia was diagnosed

**Supplementary Figure 1.** Kaplan Meier survival curve for the outcome of mortality by data source where dementia first recorded.

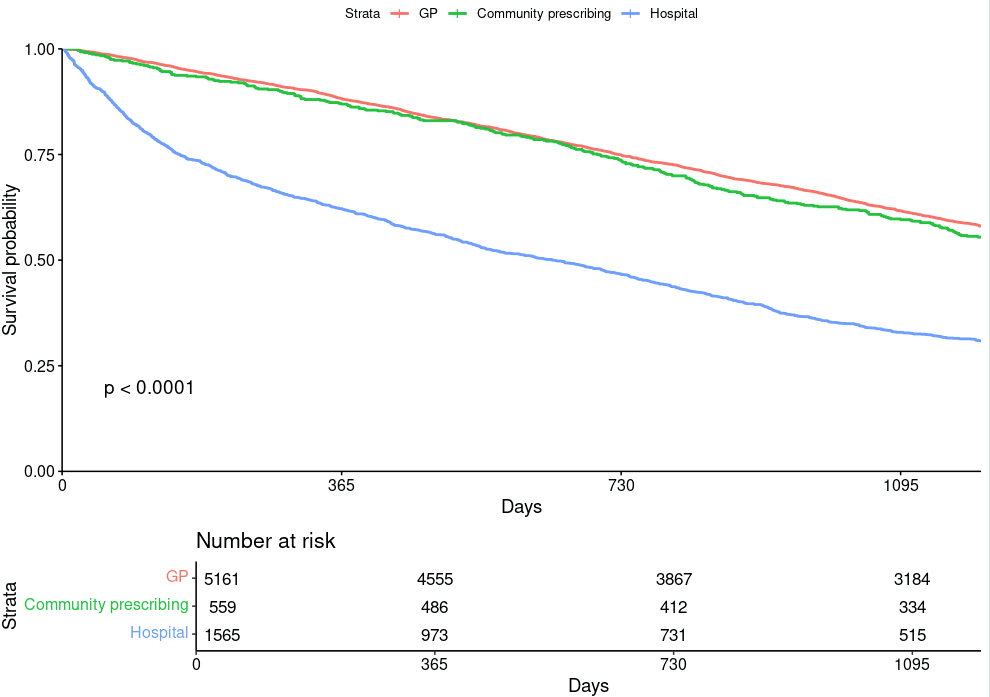

**Appendix 1.** Read version 2 and ICD-10 code lists.

**Dementia**

Source: HDR UK Phenotype library, available at <https://phenotypes.healthdatagateway.org/phenotypes/PH148/version/296/detail/>

| **code** | **description** | **coding_system** |
| --- | --- | --- |
| **Eu01000** | [X]Vascular dementia of acute onset | Read codes v2 |
| **1461.00** | H/O: dementia | Read codes v2 |
| **E00y.11** | Presbyophrenic psychosis | Read codes v2 |
| **F110.00** | Alzheimer's disease | Read codes v2 |
| **E001.00** | Presenile dementia | Read codes v2 |
| **F110100** | Alzheimer's disease with late onset | Read codes v2 |
| **Eu01y00** | [X]Other vascular dementia | Read codes v2 |
| **Eu00000** | [X]Dementia in Alzheimer's disease with early onset | Read codes v2 |
| **E00y.00** | Other senile and presenile organic psychoses | Read codes v2 |
| **E00..00** | Senile and presenile organic psychotic conditions | Read codes v2 |
| **E001z00** | Presenile dementia NOS | Read codes v2 |
| **9Ou2.00** | Dementia monitoring second letter | Read codes v2 |
| **9hD0.00** | Excepted from dementia quality indicators: Patient unsuitabl | Read codes v2 |
| **Eu01111** | [X]Predominantly cortical dementia | Read codes v2 |
| **Eu00200** | [X]Dementia in Alzheimer's dis, atypical or mixed type | Read codes v2 |
| **Eu00z11** | [X]Alzheimer's dementia unspec | Read codes v2 |
| **E003.00** | Senile dementia with delirium | Read codes v2 |
| **E041.00** | Dementia in conditions EC | Read codes v2 |
| **E004.00** | Arteriosclerotic dementia | Read codes v2 |
| **E004100** | Arteriosclerotic dementia with delirium | Read codes v2 |
| **E002100** | Senile dementia with depression | Read codes v2 |
| **Eu01.11** | [X]Arteriosclerotic dementia | Read codes v2 |
| **E002z00** | Senile dementia with depressive or paranoid features NOS | Read codes v2 |
| **66h..00** | Dementia monitoring | Read codes v2 |
| **Eu02z11** | [X] Presenile dementia NOS | Read codes v2 |
| **Eu01.00** | [X]Vascular dementia | Read codes v2 |
| **E002.00** | Senile dementia with depressive or paranoid features | Read codes v2 |
| **8CMZ.00** | Dementia care plan | Read codes v2 |
| **Eu02z14** | [X] Senile dementia NOS | Read codes v2 |
| **E001200** | Presenile dementia with paranoia | Read codes v2 |
| **Eu01300** | [X]Mixed cortical and subcortical vascular dementia | Read codes v2 |
| **Eu01z00** | [X]Vascular dementia, unspecified | Read codes v2 |
| **Eu02z15** | [X] Senile psychosis NOS | Read codes v2 |
| **E004200** | Arteriosclerotic dementia with paranoia | Read codes v2 |
| **E004300** | Arteriosclerotic dementia with depression | Read codes v2 |
| **6AB..00** | Dementia annual review | Read codes v2 |
| **Eu00011** | [X]Presenile dementia,Alzheimer's type | Read codes v2 |
| **Eu00111** | [X]Alzheimer's disease type 1 | Read codes v2 |
| **E004.11** | Multi infarct dementia | Read codes v2 |
| **Eu02z12** | [X] Presenile psychosis NOS | Read codes v2 |
| **Eu00012** | [X]Primary degen dementia, Alzheimer's type, presenile onset | Read codes v2 |
| **Eu04100** | [X]Delirium superimposed on dementia | Read codes v2 |
| **E00..12** | Senile/presenile dementia | Read codes v2 |
| **E004z00** | Arteriosclerotic dementia NOS | Read codes v2 |
| **Eu02z13** | [X] Primary degenerative dementia NOS | Read codes v2 |
| **F110000** | Alzheimer's disease with early onset | Read codes v2 |
| **E002000** | Senile dementia with paranoia | Read codes v2 |
| **Eu00.00** | [X]Dementia in Alzheimer's disease | Read codes v2 |
| **Eu00113** | [X]Primary degen dementia of Alzheimer's type, senile onset | Read codes v2 |
| **Eu00z00** | [X]Dementia in Alzheimer's disease, unspecified | Read codes v2 |
| **9Ou4.00** | Dementia monitoring verbal invite | Read codes v2 |
| **9hD1.00** | Excepted from dementia quality indicators: Informed dissent | Read codes v2 |
| **9Ou5.00** | Dementia monitoring telephone invite | Read codes v2 |
| **9Ou3.00** | Dementia monitoring third letter | Read codes v2 |
| **9Ou1.00** | Dementia monitoring first letter | Read codes v2 |
| **Eu02z00** | [X] Unspecified dementia | Read codes v2 |
| **Eu00112** | [X]Senile dementia,Alzheimer's type | Read codes v2 |
| **Eu01100** | [X]Multi-infarct dementia | Read codes v2 |
| **9hD..00** | Exception reporting: dementia quality indicators | Read codes v2 |
| **E001000** | Uncomplicated presenile dementia | Read codes v2 |
| **E001300** | Presenile dementia with depression | Read codes v2 |
| **Eu01200** | [X]Subcortical vascular dementia | Read codes v2 |
| **ZS7C500** | Language disorder of dementia | Read codes v2 |
| **E00z.00** | Senile or presenile psychoses NOS | Read codes v2 |
| **E00..11** | Senile dementia | Read codes v2 |
| **Fyu3000** | [X]Other Alzheimer's disease | Read codes v2 |
| **Eu00100** | [X]Dementia in Alzheimer's disease with late onset | Read codes v2 |
| **E001100** | Presenile dementia with delirium | Read codes v2 |
| **E004000** | Uncomplicated arteriosclerotic dementia | Read codes v2 |
| **9Ou..00** | Dementia monitoring administration | Read codes v2 |
| **Eu02z16** | [X] Senile dementia, depressed or paranoid type | Read codes v2 |
| **Eu00013** | [X]Alzheimer's disease type 2 | Read codes v2 |
| **E000.00** | Uncomplicated senile dementia | Read codes v2 |
| **F05.1** | Delirium superimposed on dementia | ICD10 codes |
| **G30** | Alzheimer's disease | ICD10 codes |
| **F03** | Unspecified dementia | ICD10 codes |
| **F01** | Vascular dementia | ICD10 codes |
| **F00** | Dementia in Alzheimer's disease | ICD10 codes |

**Memory problems**

eFI version 1

| Memory & cognitive problems | 1461. | H/O: dementia |
| --- | --- | --- |
| Memory & cognitive problems | 1B1A. | Memory loss - amnesia |
| Memory & cognitive problems | 1S21. | Disturb of mem for ord events |
| Memory & cognitive problems | 2841. | Confused |
| Memory & cognitive problems | 28E.. | Cognitive decline |
| Memory & cognitive problems | 3A10. | Memory: own age not known |
| Memory & cognitive problems | 3A20. | Memory: present time not known |
| Memory & cognitive problems | 3A30. | Memory: present place not knwn |
| Memory & cognitive problems | 3A40. | Memory: present year not known |
| Memory & cognitive problems | 3A50. | Memory: own DOB not known |
| Memory & cognitive problems | 3A60. | Memory: present month not knwn |
| Memory & cognitive problems | 3A70. | Memory: important event not kn |
| Memory & cognitive problems | 3A80. | Memory: import.person not knwn |
| Memory & cognitive problems | 3A91. | Memory: count down unsuccess. |
| Memory & cognitive problems | 3AA1. | Memory: address recall unsucc. |
| Memory & cognitive problems | 3AE.. | GDS: assess prim deg dement |
| Memory & cognitive problems | 66h.. | Dementia monitoring |
| Memory & cognitive problems | 6AB.. | Dementia annual review |
| Memory & cognitive problems | 8HTY. | Referral to memory clinic |
| Memory & cognitive problems | 9NdL. | Lacks capacity consnt MCA 2005 |
| Memory & cognitive problems | 9Nk1. | Seen in memory clinic |
| Memory & cognitive problems | 9Ou.. | Dementia monitoring admin. |
| Memory & cognitive problems | 9Ou2. | Dementia monitoring 2nd letter |
| Memory & cognitive problems | 9Ou3. | Dementia monitoring 3rd letter |
| Memory & cognitive problems | 9Ou4. | Dementia monitor verbal invite |
| Memory & cognitive problems | 9Ou5. | Dementia monitor phone invite |
| Memory & cognitive problems | 9hD.. | Excep report: demen qual indic |
| Memory & cognitive problems | 9hD0. | Exc demen qual ind: Pat unsuit |
| Memory & cognitive problems | 9hD1. | Exc demen qual ind: Inform dis |
| Memory & cognitive problems | E00.. | Senile/presenile organic psych |
| Memory & cognitive problems | E000. | Senile dementia-uncomplicated |
| Memory & cognitive problems | E001. | Presenile dementia |
| Memory & cognitive problems | E0010 | Presenile dementia - uncomplic |
| Memory & cognitive problems | E0011 | Presenile dementia + delirium |
| Memory & cognitive problems | E0012 | Presenile dementia + paranoia |
| Memory & cognitive problems | E0013 | Presenile dementia+depression |
| Memory & cognitive problems | E001z | Presenile dementia NOS |
| Memory & cognitive problems | E002. | Sen.dement.-depressed/paranoid |
| Memory & cognitive problems | E0020 | Senile dementia + paranoia |
| Memory & cognitive problems | E0021 | Senile dementia + depression |
| Memory & cognitive problems | E002z | Sen.dement.-depr./paranoid NOS |
| Memory & cognitive problems | E003. | Senile dementia + delirium |
| Memory & cognitive problems | E004. | Arteriosclerotic dementia |
| Memory & cognitive problems | E0040 | Arterioscl.dementia-uncomplic. |
| Memory & cognitive problems | E0041 | Arterioscl.dementia+delirium |
| Memory & cognitive problems | E0042 | Arterioscl.dementia+paranoia |
| Memory & cognitive problems | E0043 | Arterioscl.dementia+depression |
| Memory & cognitive problems | E004z | Arteriosclerotic dementia NOS |
| Memory & cognitive problems | E012. | Other alcoholic dementia |
| Memory & cognitive problems | E041. | Dementia in conditions EC |
| Memory & cognitive problems | E2A10 | Mild memory disturbance |
| Memory & cognitive problems | E2A11 | Organic memory impairment |
| Memory & cognitive problems | Eu00. | [X]Dementia in Alzheimer's |
| Memory & cognitive problems | Eu000 | [X]Early onset Alzheim dement |
| Memory & cognitive problems | Eu001 | [X]Late onset Alzheim dementia |
| Memory & cognitive problems | Eu002 | [X]Atypical/mixed Alzheimer's |
| Memory & cognitive problems | Eu00z | [X]Alzheimer's disease unspec |
| Memory & cognitive problems | Eu01. | [X]Vascular dementia |
| Memory & cognitive problems | Eu010 | [X]Vascular dement acute onset |
| Memory & cognitive problems | Eu011 | [X]Multi-infarct dementia |
| Memory & cognitive problems | Eu012 | [X]Subcortical vascular dement |
| Memory & cognitive problems | Eu013 | [X]Mix cort/subcor vasc dement |
| Memory & cognitive problems | Eu01y | [X]Other vascular dementia |
| Memory & cognitive problems | Eu01z | [X]Vascular dementia unspecif |
| Memory & cognitive problems | Eu02. | [X]Dementia in disease EC |
| Memory & cognitive problems | Eu020 | [X]Dementia in Pick's disease |
| Memory & cognitive problems | Eu021 | [X]Dement in Creutzfeld-Jakob |
| Memory & cognitive problems | Eu022 | [X]Dementia in Huntington's |
| Memory & cognitive problems | Eu023 | [X]Dementia in Parkinson's |
| Memory & cognitive problems | Eu024 | [X]Dementia in HIV disease |
| Memory & cognitive problems | Eu025 | [X]Lewy body dementia |
| Memory & cognitive problems | Eu02y | [X]Dement,oth sp dis cl elsewh |
| Memory & cognitive problems | Eu02z | [X] Unspecified dementia |
| Memory & cognitive problems | Eu041 | [X]Delirium superimp dementia |
| Memory & cognitive problems | Eu057 | [X]Mild cognitive disorder |
| Memory & cognitive problems | F110. | Alzheimer's disease |
| Memory & cognitive problems | F1100 | Alzheimer dis wth early onset |
| Memory & cognitive problems | F1101 | Alzheimer's dis wth late onset |
| Memory & cognitive problems | F116. | Lewy body disease |
| Memory & cognitive problems | F21y2 | Binswanger's disease |
| Memory & cognitive problems | R00z0 | [D]Amnesia (retrograde) |
